# UK prevalence of underlying conditions which increase the risk of severe COVID-19 disease: a point prevalence study using electronic health records

**DOI:** 10.1101/2020.08.24.20179192

**Authors:** Jemma L Walker, Daniel J Grint, Helen Strongman, Rosalind M Eggo, Maria Peppa, Caroline Minassian, Kathryn E Mansfield, Christopher T Rentsch, Ian J Douglas, Rohini Mathur, Angel YS Wong, Jennifer K Quint, Nick Andrews, Jamie Lopez Bernal, J Anthony Scott, Mary Ramsay, Liam Smeeth, Helen McDonald

**Affiliations:** NIHR Health Protection Research Unit (HPRU) in Immunisation, London, UK; London School of Hygiene & Tropical Medicine, Keppel Street, London, WC1E 7HT, UK; Public Health England, 61 Colindale Ave, London NW9 5EQ, UK; Imperial College London, South Kensington, London SW7 2BU, UK

## Abstract

**Background:** This study aimed to describe the population at risk of severe COVID-19 due to underlying health conditions across the United Kingdom in 2019.

**Methods:** We used anonymised electronic health records from the Clinical Practice Research Datalink GOLD to describe the point prevalence on 5 March 2019 of the at-risk population following national guidance. Prevalence for any risk condition and for each individual condition is given overall and stratified by age and region. We repeated the analysis on 5 March 2014 for full regional representation and to describe prevalence of underlying health conditions in pregnancy. We additionally described the population of cancer survivors, and assessed the value of linked secondary care records for ascertaining COVID-19 at-risk status.

**Findings:** On 5 March 2019, 24·4% of the UK population were at risk due to a record of at least one underlying health condition, including 8·3% of school-aged children, 19·6% of working-aged adults, and 66·2% of individuals aged 70 years or more. 7·1% of the population had multimorbidity. The size of the at-risk population was stable over time comparing 2014 to 2019, despite increases in chronic liver disease and diabetes and decreases in chronic kidney disease and current asthma. Separately, 1·6% of the population had a new diagnosis of cancer in the past five years.

**Interpretation:** The population at risk of severe COVID-19 (aged ≥70 years, or with an underlying health condition) comprises 18.5 million individuals in the UK, including a considerable proportion of school-aged and working-aged individuals.

**Funding:** NIHR HPRU in Immunisation

**Research in context:** *Evidence before this study:* We searched Pubmed for peer-reviewed articles, preprints, and research reports on the size and distribution of the population at risk of severe COVID. We used the terms (1) risk factor or co-morbidity or similar (2) COVID or SARS or similar and (3) prevalence to search for studies aiming to quantify the COVID-19 at-risk UK population published in the previous year to 19 July 2020, with no language restrictions. We found one study which modelled prevalence of risk factors based on the Global Burden of Disease (which included the UK) and one study which estimated that 8.4 million individuals aged ≥30 years in the UK were at risk based on prevalence of a subset of relevant conditions in England. There were no studies which described the complete COVID-19 at-risk population across the UK.

*Added value of this study:* We used a large, nationally-representative dataset based on electronic health records to estimate prevalence of increased risk of severe COVID-19 across the United Kingdom, including all conditions in national guidance. We stratified by age, sex and region to enable regionally-tailored prediction of COVID-19-related healthcare burden and interventions to reduce transmission of infection, and planning and modelling of vaccination of the at-risk population. We also quantified the value of linked secondary care records to supplement primary care records.

*Implications of all the available evidence:* Individuals at moderate or high risk of severe COVID-19 according to current national guidance (aged ≥70 years, or with a specified underlying health condition) comprise 18·5 million individuals in the United Kingdom, rather than the 8.43 million previously estimated. The 8·3% of school-aged children and 19·6% of working-aged adults considered at-risk according to national guidance emphasises the need to consider younger at-risk individuals in shielding policies and when re-opening schools and workplaces, but also supports prioritising vaccination based on age and condition-specific mortality risk, rather than targeting all individuals with underlying conditions, who form a large population even among younger age groups. Among individuals aged ≥70 years, 66·2% had at least one underlying health condition, suggesting an age-targeted approach to vaccination may efficiently target individuals at risk of severe COVID-19. These national estimates broadly support the use of Global Burden of Disease modelled estimates and age-targeted vaccination strategies in other countries.

## Introduction

People with underlying health conditions account for the majority of COVID-19-related hospital and intensive care admissions, and are at increased risk of death from COVID-19 compared to the general population of the same age.^1-4^ Prevalence of many conditions increases with age, which is also an independent risk factor for COVID-19 mortality.^3, 4^

Characterising the population at risk of severe COVID-19 is vital for effective policy and planning in response to the COVID-19 pandemic.^5^ Age-and region-specific prevalence of at-risk groups are key to predicting mortality and managing pressure on hospital inpatient and intensive care services across the country. Numbers of school-aged children and working-aged adults at risk are important for re-opening local schools and workplaces. Vaccination planning requires at-risk population size for vaccine numbers, and age and regional distribution for modelling impact on regional transmission, since vaccine response typically decreases with older age.^6^

Modelling based on the Global Burden of Disease (GBD) study suggests that approximately one in five individuals worldwide have a health condition that increases risk of COVID-19.^7^ National prevalence studies of COVID-19 at-risk groups are rare. Large household surveys suggest that a third of adults in the United States, and between a third and a half of adults in Brazil, have at least one risk factor for COVID-19 (based on age ≥65 years, or underlying health conditions for younger adults).^8, 9^ A previous study estimated that at least 8.4 million individuals in the UK were at risk, but included only a subset of relevant health conditions.^10^ Universal healthcare with an electronic health records system offers an opportunity for precise and representative estimation of at-risk prevalence in the UK which may aid in interpretation of GBD-based estimates elsewhere, and support UK policy.

This study aimed to quantify the size, composition, and distribution of the population at risk of severe COVID-19 across the UK in March 2019, using electronic health records to define at-risk status based on all underlying conditions in national guidance.

## Methods

### Data sources

We conducted a point prevalence study among the UK general population using the Clinical Practice Research Datalink (CPRD) GOLD dataset, an anonymised sample of electronic health records from primary care practices across the UK.^11^ The dataset includes diagnoses recorded using Read codes, primary care prescribing, and results of tests ordered in primary care. Data validity has been shown to be high.^12^

Secondary care (hospital) data linkage is available for approximately 75% of CPRD GOLD-registered individuals in England, based on practice-level consent. For patients admitted to hospital, the Hospital Episode Statistics Admitted Patient Care dataset records diagnoses using International Classification of Diseases ICD-10 codes, and procedures such as chemotherapy using Classification of Interventions and Procedures OPCS-4 codes.^13^

The CPRD Pregnancy Register uses validated algorithms, combining information across the primary care record such as antenatal scans, expected delivery dates, and deliveries, terminations and miscarriage records, to date and characterise pregnancies in CPRD GOLD.^14^

### Index dates

Our primary analysis index date was 5 March 2019 for up-to-date national prevalence estimates. CPRD GOLD coverage peaked in 2014, when it included approximately 7% of the UK population: by 2019 the dataset was smaller and did not cover all regions in England. As a secondary analysis, point prevalence estimates were repeated for 5 March 2014 for greater power and full regional representation across England.

Pregnancy was described for the index date of 5 March 2014 only, not 5 March 2019, since the latest Pregnancy Register update was in February 2018.

### Study population

The study population comprised individuals aged 2–100 years active in CPRD GOLD, with at least one year’s prior registration to allow recording of underlying conditions.^15^ Eligibility started on the latest of: 1 January 2019, second birthday, a year after registration, or practice meeting CPRD quality standards. Eligibility ended at the earliest of: 5 March 2019, hundredth birthday, death, leaving the practice, or last data collection from the practice. Individuals with any time eligible between 1 January and 5 March were included in the main analysis of point prevalence on 5 March to increase study power, with a sensitivity analysis limited to individuals active in the dataset on 5 March 2019.

For pregnancy, the study population comprised women aged 11–49 years. As pregnancy is transient, women were required to be active in the dataset on 5 March 2014, rather than any time between 1 January and 5 March 2014.

### Definition of at-risk population

In national guidance, all individuals aged ≥70 years are considered at moderate risk (**Box 1**).^1^ Since age-specific population estimates are readily available, the primary analysis for this study defined at-risk status based on underlying health conditions alone, rather than age. An additional analysis estimated the size of the at-risk population including all individuals aged ≥70 years.

We defined the COVID-19 at-risk population as individuals with *at least one* underlying health condition conferring moderate or high risk of severe COVID-19 according to national guidance (**Box 1**). Namely: any history of chronic respiratory disease (excluding asthma), heart disease, kidney disease, neurological conditions such as multiple sclerosis, diabetes mellitus; or current asthma, severe obesity, or immunosuppression; assessed on the index date.^1^

Underlying conditions were defined using diagnoses, height and weight measurements, test results, and prescriptions recorded in primary care for the main analysis. Pregnancy status was ascertained from the CPRD Pregnancy Register (**Supplementary Table 1**). Individuals with no recorded body mass index were included in the analysis, categorised as having no evidence of severe obesity. For analysis using linked secondary care data, diagnoses and procedures recorded in secondary care were additionally ascertained from ICD-10 and OCPS-4 codes respectively.

**Table 1:**
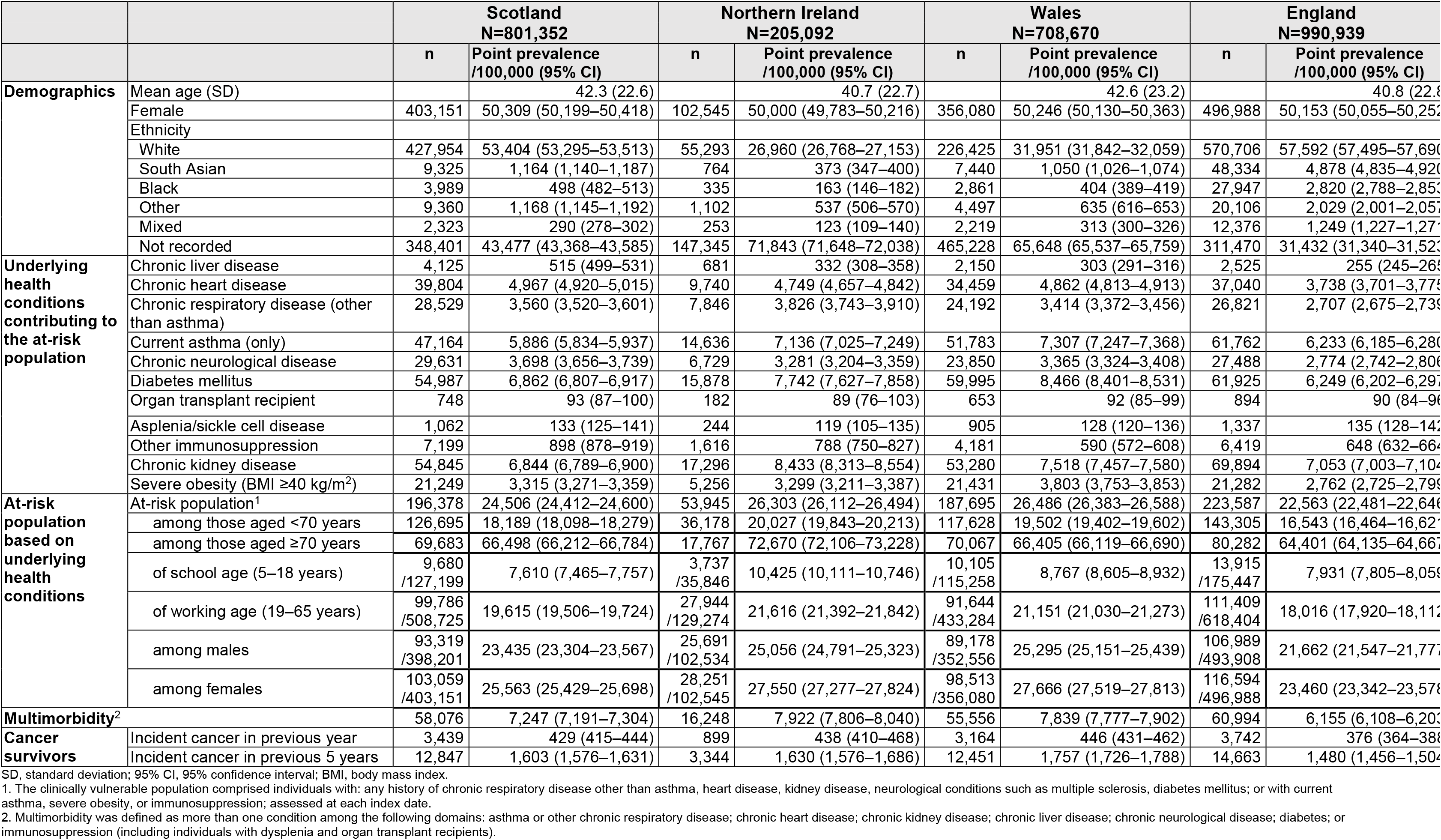
Study population characteristics and point prevalence of the COVID-19 at-risk population on 5 March 2019 in the United Kingdom using standalone primary care data, N=2,706,053

Cancer survivors have an increased risk of COVID-19 mortality but non-haematological cancer survivors are only included in current COVID-19 guidance if receiving immunosuppressing treatment (**Box 1**).^3^ Separately to the study at-risk definition we described prevalence of any new cancer diagnosis in the past one and five years.^16^

### Statistical Analysis

Point prevalence estimates of the at-risk population and each underlying condition on 5 March 2019 were calculated per 100,000 with binomial exact 95% confidence intervals, for each nation in the UK.

The at-risk population prevalence was stratified by sex and age, categorised in 5-year bands except 2–9 years and 90–99 years. Prevalence estimates for the at-risk population and each condition were stratified by age and region, separately and in combination. Prevalence values with fewer than five individuals were suppressed to preserve confidentiality.

For additional analysis estimating the size of the at-risk population including all individuals aged ≥70 years, the at-risk prevalence among individuals aged 2–69 years was age-standardised in 5-year bands, and added to the population aged ≥70 years, using mid-2019 national population estimates.^17^

Comparison of prevalence in 2014 to 2019 was stratified by region to account for the change in regional representation of the dataset over time.

The point prevalence of pregnancy and underlying health conditions was estimated among women aged 11-49 years on 5 March 2014.

Prevalence estimates with and without linked secondary care records were compared among individuals at practices in England which had consented to data linkage.

#### Sensitivity analyses

CPRD GOLD was nationally representative by age and sex in March 2011.^11^ To update this assessment, the 2019 study population was compared to mid-2019 national population estimates, and 2019 at-risk prevalence estimates directly age-standardised in five-year bands using mid-2019 population estimates for each nation.^17^

The main analysis included individuals active in the dataset at any point between 1 January and 5 March 2019. Individuals who left CPRD between 1 January and 5 March would not subsequently have had new diagnoses recorded, which could underestimate point prevalence on 5 March. As a sensitivity analysis, at-risk prevalence was estimated restricted to individuals active in CPRD on 5 March 2019.

All analysis was conducted using STATA 16 MP.

### Ethics approval

Approval was received from the Independent Scientific Advisory Committee of the Medicines and Healthcare Products Regulatory Agency (ISAC number: 20_062A) and the Ethics Committee of the London School of Hygiene and Tropical Medicine (reference 21851). The ISAC protocol was made available to reviewers.

## Results

### Characteristics of the study population

The 2019 study population included 2,706,053 individuals: 990,939 (36·6%) in England, 801,352 in Scotland, 708,670 in Wales and 205,092 in Northern Ireland (**Table 1**). Approximately half (50·2%) were female. The study included 359,412 individuals (13·3%) aged ≥70 years. There was some over-representation of 40–59-year-olds compared to mid-2019 national population estimates for all four countries (**Figure 1**).

**Figure 1:**
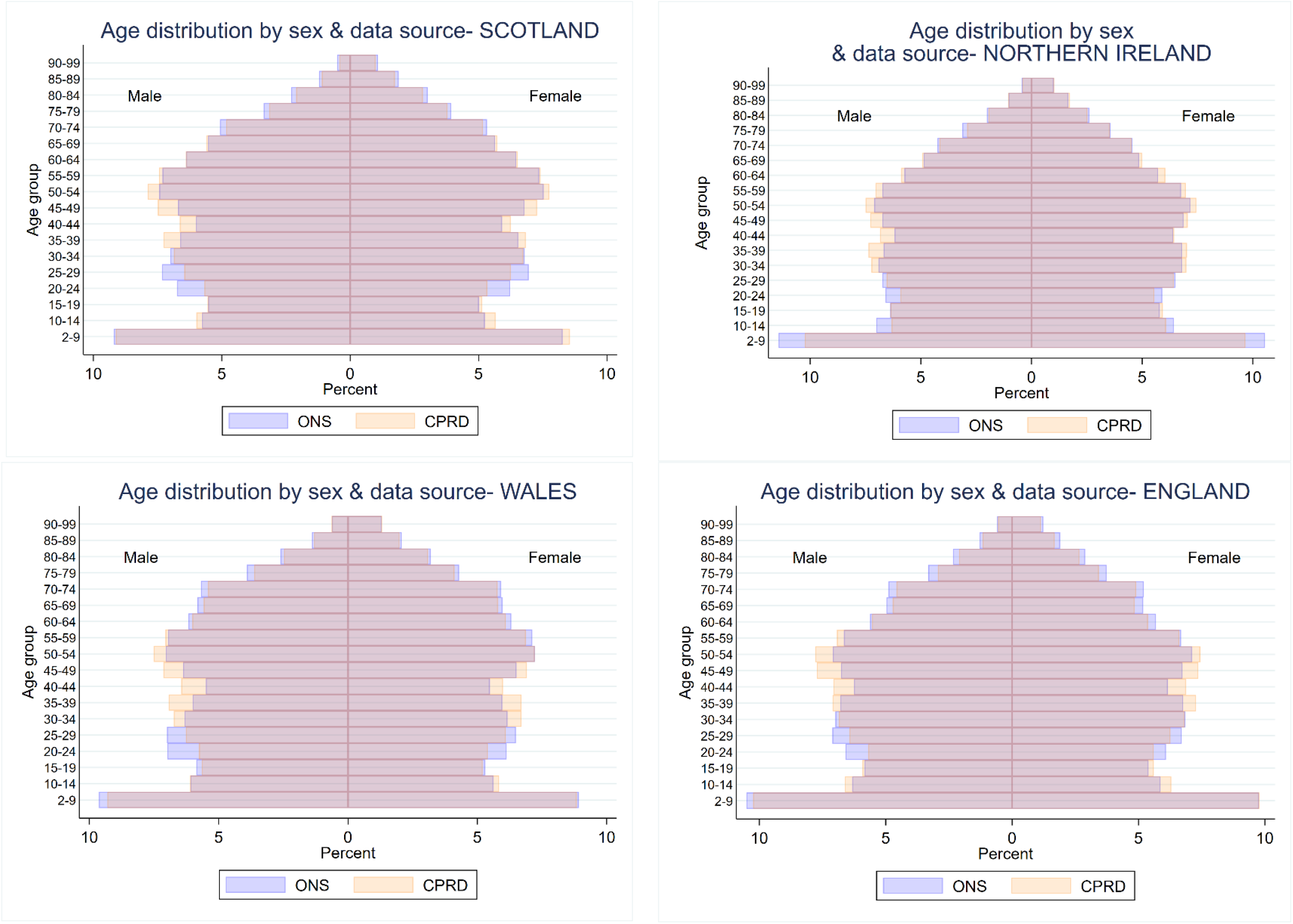
Age and sex distributions of the CPRD GOLD study population on 5 March 2019 (N=2,706,053) compared to mid-2019 national population estimates^17^

In 2014, the dataset included 4,730,254 individuals: 2,980,402 (63·0%) in England, 810,169 (17·1%) in Scotland, 730,563 (15·4%) in Wales and 209,120 (4·4%) in Northern Ireland (Supplementary Table 2). Age and sex distributions were similar to 2019, with 50·3% female and 12·7% aged ≥70 years.

### COVID-19 at-risk population

On 5 March 2019, 24·4% (95% CI 24·4-24·5) of the study population were at risk of severe COVID-19 due to underlying health conditions. National at-risk prevalence ranged from 22·6% in England to 26·5% in Wales (**Table 1**).

In a secondary analysis, the number of at-risk individuals based on current guidance including all individuals aged ≥70 years was estimated at 18.5 million across the UK, of whom 9·53 million (95% CI 9·52-9·53) were aged <70 years (**Table 2**).

**Table 2:** Estimated size of the at-risk population in 2019 based on either age (≥ 70 years) or underlying health conditions among individuals aged 2 to 69 years

|  | Age 2 to 69 years |  |  | Age 70 years or older | Estimated total number of individuals in the at-risk population |
| --- | --- | --- | --- | --- | --- |
|  | Age-standardised prevalence of at least one underlying health condition /100,000 (95% CI) | Office for National Statistics mid-2019 population estimate | Estimated number of individuals aged 2–69 years with at least one underlying health condition (95% CI) | Office for National Statistics mid-2019 population estimate |  |
| **Scotland** | 18,046 (17,956–18,137) | 4,615,093 | 832,848 (828,694–837,019) | 744,701 | 1,577,549 |
| **Northern Ireland** | 19,672 (19,490–19,855) | 1,622,687 | 319,213 (316,266–322,182) | 224,851 | 544,064 |
| **Wales** | 19,519 (19,418–19,619) | 2,609,759 | 509,389 (506,776–512,011) | 480,234 | 989,623 |
| **England** | 16,573 (16,495–16,652) | 47,467,071 | 7,866,870 (7,829,660–7,904,217) | 7,556,976 | 15,423,846 |

95% CI, 95% confidence interval

#### Composition by underlying health conditions

The commonest conditions across the UK were chronic kidney disease (7·2%), diabetes mellitus (7·1%), asthma (6·5%) and chronic heart disease (4·5%). Prevalence of each condition varied nationally, with chronic liver disease notably commoner in Scotland (**Table 1**). Multimorbidity was common, ranging from 6·2% in England to 7·9% in Northern Ireland: 7·1% across the UK.

#### Variation by age

The proportion of at-risk individuals increased gradually with age from 5·1% of children aged 2-9 years to a peak at 79·4% of those aged 85-89 years in England before declining at older ages (**Figure 2**). Similar age distributions were seen in each nation, and for each condition except current asthma, which peaked at age 10-14 years (**Figure 2**).

**Figure 2:**
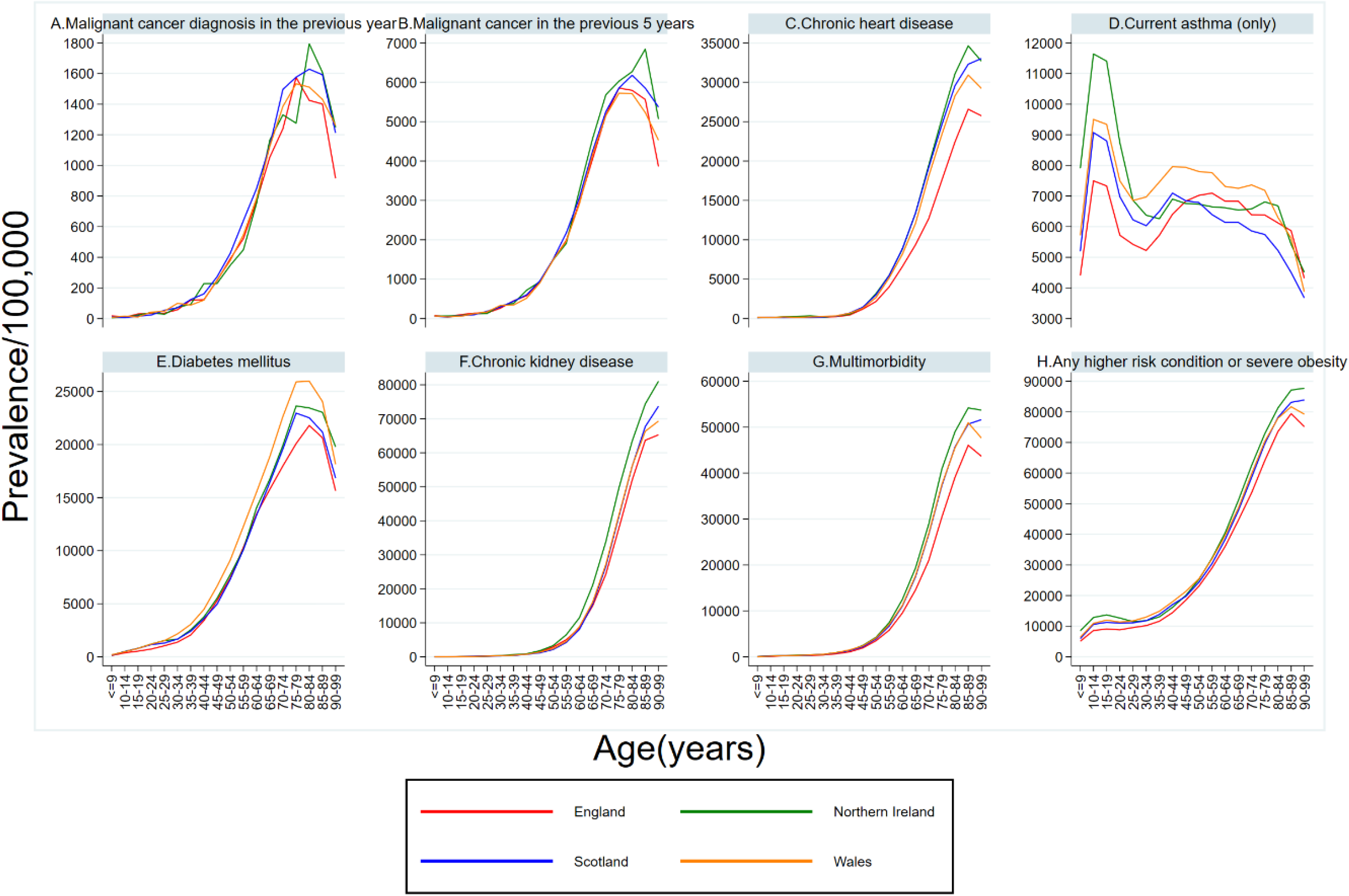
Age distributions of the at-risk population and common contributing underlying health conditions on 5 March 2019, N=2,706,053

The at-risk population comprised 18·1% of individuals aged <70 years (including 8·3% of school-aged children and 19·6% of working aged adults) and 66·2% of individuals aged ≥70 years across the UK (**Table 1**).

#### Variation by sex

Overall, a higher proportion of women than men were at risk (**Table 1**), but the association varied with age, and men were more likely than women to be at risk from age 55 years upwards (**Figure 3, Supplementary Table 3**).

**Figure 3:**
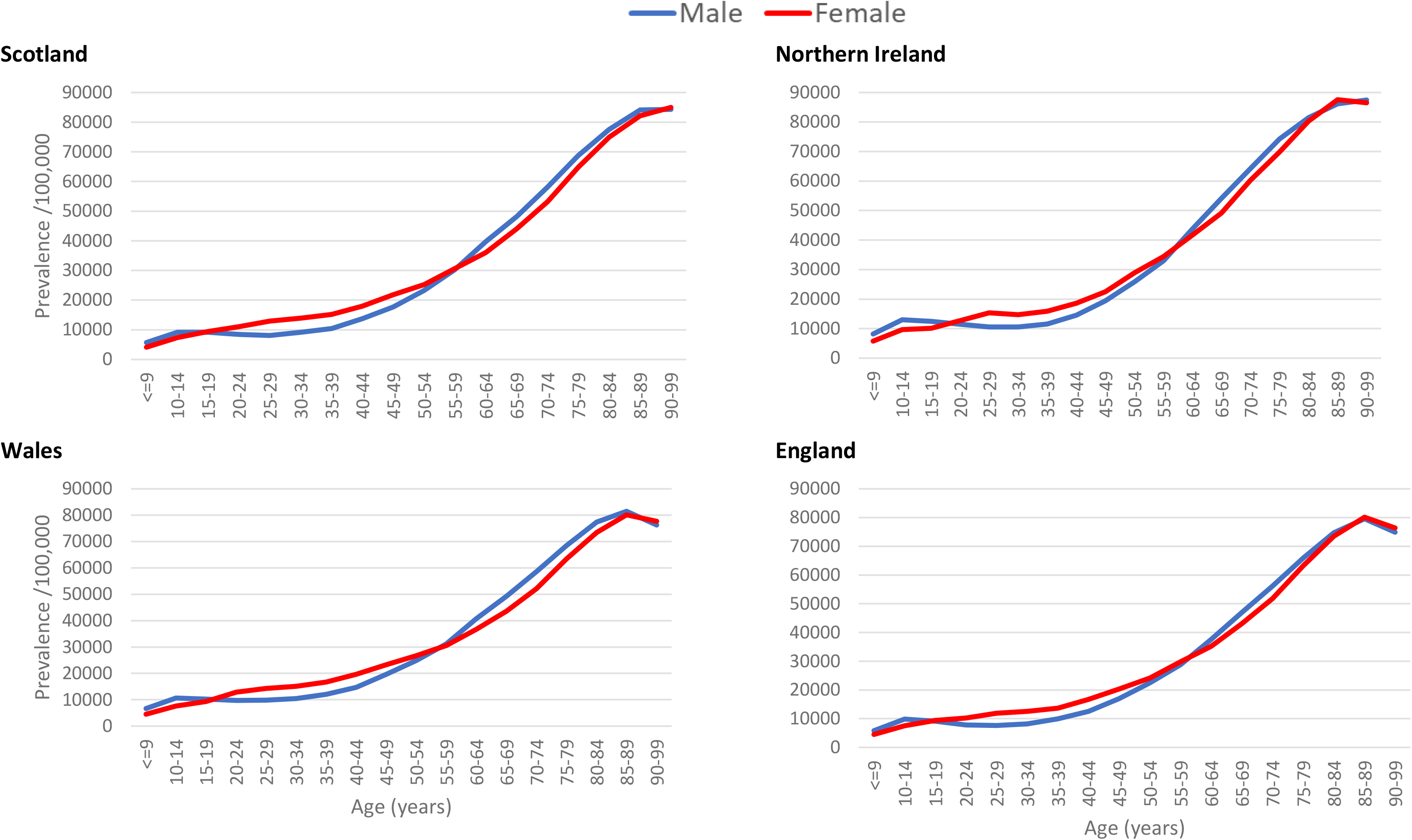
Prevalence of the at-risk population by age and sex across the United Kingdom on 5 March 2019, N=2,706,053

#### Variation by region

No individuals from the North East or East Midlands regions of England were included in 2019, whereas all regions were represented in 2014. London had the lowest proportion of the population considered at risk in both 2014 and 2019 (**Figure 4**). The East of England, South Central and South East also had lower prevalence of at-risk individuals than Midlands or Northern regions in both 2014 and 2019. Regional patterns varied between underlying conditions (**Supplementary Figure 1**).

**Figure 4:**
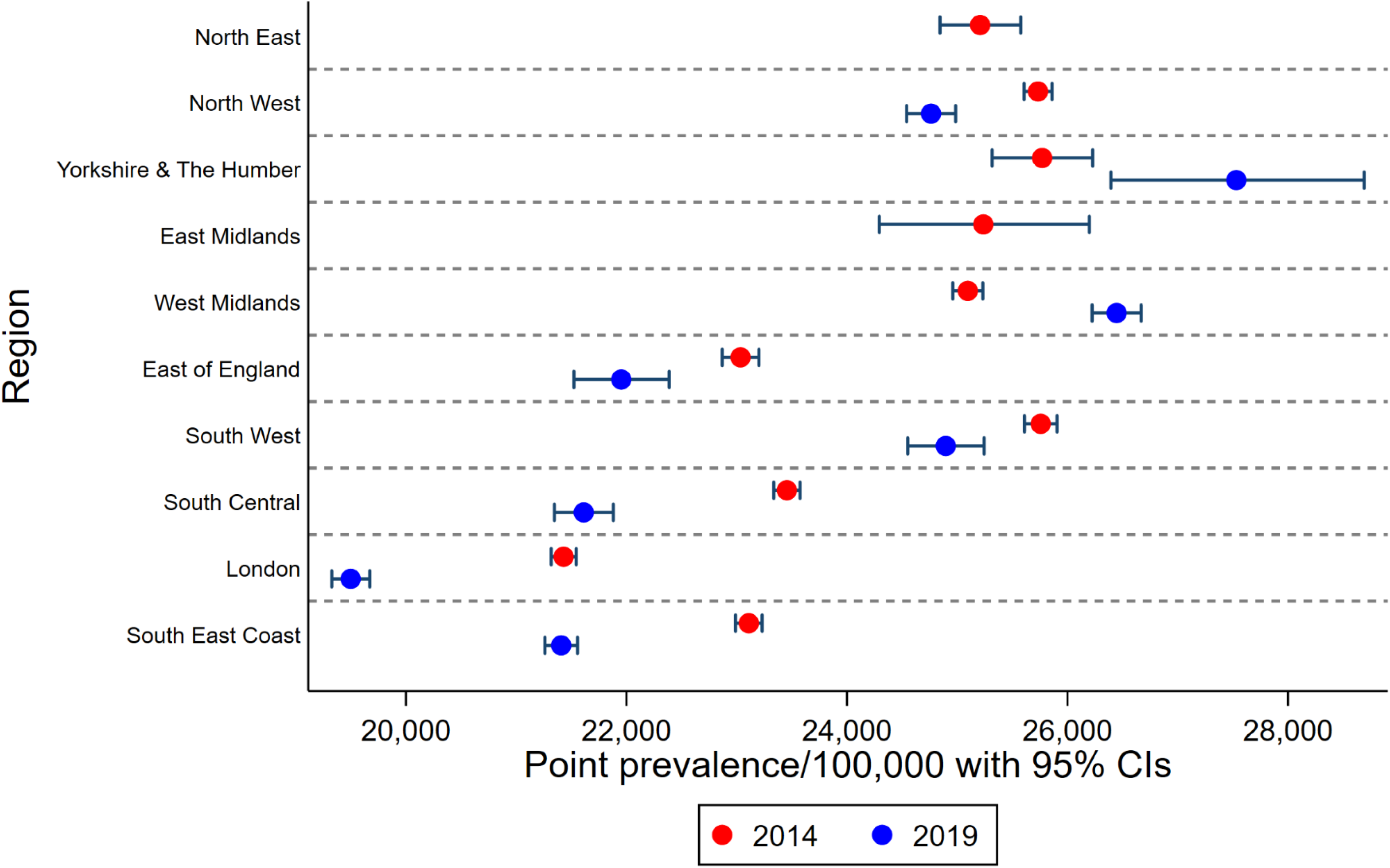
Point prevalence of the at-risk population by region in England comparing 5 March 2014 (N=4,730,254) and 5 March 2019 (N=2,706,053) 2019 study population did not include any individuals in the North East or East Midlands regions; x-axis scale starts at 20,000/100,000.

Prevalence estimates of the at-risk population and each condition stratified by age and region (separately and combined) on 5 March 2014 and 2019 are here: https://doi.org/10.17037/DATA.00001833

#### Differences between 2014 and 2019 prevalence estimates

Compared to 2014, at-risk prevalence estimates in 2019 were 0·8% higher in Northern Ireland but lower in Scotland (−0·5%), Wales (−0·8%), and England (−1·3%). When stratified by region within England (**Figure 4**), at-risk prevalence increased from 2014 to 2019 for Yorkshire and the Humber and the West Midlands, and decreased in all other regions (excluding the North East and East Midlands, which were unavailable in the 2019 dataset), but no changes exceeded 1·9% difference.

For underlying conditions, absolute changes in UK prevalence estimates from 2014 to 2019 ranged from a -0·9% decrease in chronic kidney disease to a 0·7% increase in diabetes mellitus (**Supplementary Figure 1**). The biggest relative increases were for chronic liver disease (+32·3% from 2014 to 2019), diabetes (+11·5%) and chronic respiratory disease other than asthma (+11·4%). The largest relative falls were for chronic kidney disease (−10·6%) and current asthma (−6·0%).

#### Cancer survivors

On 5 March 2019, 0·4% of the UK had incident cancer recorded within the previous year and 1·6% within the previous five years (**Table 1**).

#### Pregnancy

Among women aged 11–49 years on 5 March 2014, 2·1% were pregnant, of whom 12·9% had a recorded health condition, compared to 14·5% of non-pregnant women (**Table 3**).

**Table 3:** Prevalence of pregnancy and underlying health conditions among women aged 11 to 49 years on 5 March 2014, N=1,181,840

|  | Scotland<br>N=205,542 |  | Northern Ireland<br>N=55,559 |  | Wales<br>N=177,412 |  | England<br>N=743,327 |  |
| --- | --- | --- | --- | --- | --- | --- | --- | --- |
| **Among all women aged 11–49 years** | n | Point prevalence<br>/100,000 (95% CI) | n | Point prevalence /100,000<br>(95% CI) | n | Point prevalence<br>/100,000 (95% CI) | n | Point prevalence<br>/100,000 (95% CI) |
| Pregnant or any underlying health condition<sup>1</sup> | 34,051 | 16,566 (16,406–16,728) | 9,234 | 16,620 (16,311–16,932) | 31,140 | 17,552 (17,376–17,730) | 117,680 | 15,832 (15,749–15,915) |
| Any underlying health condition<sup>1</sup> | 31,389 | 15,271 (15,116–15,428) | 8,317 | 14,970 (14,674–15,269) | 28,733 | 16,196 (16,024–16,368) | 102,121 | 13,738 (13,660–13,817) |
| Pregnant | 3,067 | 1,492 (1,440–1,545) | 1,069 | 1,924 (1,811–2,042) | 2,834 | 1,597 (1,540–1,657) | 17,755 | 2,389 (2,354–2,424) |
| **Prevalence of underlying health conditions, stratified by pregnancy status among women aged 11–49 years** |  |  |  |  |  |  |  |  |
| Pregnant with an underlying health condition | 405 | 13,205 (12,026–14,455) | 152 | 14,219 (12,179–16,458) | 427 | 15,067 (13,769–16,458) | 2,196 | 12,368 (11,887–12,862) |
| Not pregnant, with an underlying health condition | 30,984 | 15,303 (15,146–15,460) | 8,165 | 14,984 (14,686–15,287) | 28,306 | 16,214 (16,041–16,388) | 99,925 | 13,772 (13,693–13,851) |

95% CI, 95% confidence interval
1. Underlying health conditions comprised: any history of chronic respiratory disease other than asthma, heart disease, kidney disease, neurological conditions such as multiple sclerosis, diabetes mellitus; or current asthma, severe obesity, or immunosuppression; assessed at each index date.

### Linked secondary care records

At-risk prevalence based on standalone primary care records was similar among individuals with and without eligibility for data linkage. Linked secondary care records increased the estimated prevalence of the at-risk population in England by 1·8% in both 2014 and 2019. The increase was greater among individuals <70 years than those ≥70 years.

For underlying conditions, the greatest absolute changes in prevalence estimates were for multimorbidity, which increased from 6·5% to 7·6% in 2019, and chronic heart disease, which increased from 4·0% to 5·3%. The greatest relative increase was for chronic liver disease, which nearly doubled from 0·27% to 0·53% (**Table 4**).

**Table 4:** Prevalence estimates for England with and without linked secondary care data in 2014 (N=1,802,468) and 2019 (N=744,496)

|  | Individuals in England eligible for secondary care data linkage in 2014 N=1,802,468 |  |  |  | Individuals in England eligible for secondary care data linkage in 2019 N=744,496 |  |  |  |
| --- | --- | --- | --- | --- | --- | --- | --- | --- |
|  | Standalone primary care data |  | Both primary and secondary care data |  | Standalone primary care data |  | Both primary and secondary care data |  |
|  | n | Point prevalence /100,000 (95% CI) | N | Point prevalence /100,000 (95% CI) | n | Point prevalence /100,000 (95% CI) | n | Point prevalence /100,000 (95% CI) |
| **Underlying health conditions contributing to the at-risk population** |  |  |  |  |  |  |  |  |
| Chronic liver disease | 4,499 | 250 (242–257) | 8,104 | 450 (440–459) | 2,022 | 272 (260–284) | 3,941 | 529 (513–546) |
| Chronic heart disease | 87,217 | 4,839 (4,807–4,870) | 108,711 | 6,031 (5,997–6,066) | 29,399 | 3,949 (3,905–3,993) | 39,083 | 5,250 (5,199–5,300) |
| Chronic respiratory disease | 53,358 | 2,960 (2,936–2,985) | 63,562 | 3,526 (3,500–3,553) | 21,345 | 2,867 (2,829–2,905) | 26,407 | 3,547 (3,505–3,589) |
| Current asthma (only) | 125,548 | 6,965 (6,928–7,003) | 136,702 | 7,584 (7,546–7,623) | 48,009 | 6,449 (6,393–6,505) | 53,030 | 7,123 (7,065–7,182) |
| Chronic neurological disease | 60,482 | 3,356 (3,329–3,382) | 69,932 | 3,880 (3,852–3,908) | 21,818 | 2,931 (2,892–2,969) | 26,284 | 3,530 (3,489–3,573) |
| Diabetes mellitus | 110,144 | 6,111 (6,076–6,146) | 114,149 | 6,333 (6,297–6,369) | 48,461 | 6,509 (6,453–6,566) | 50,760 | 6,818 (6,761–6,876) |
| Organ transplant recipient | 1,470 | 82 (77–86) | 1,732 | 96 (92–101) | 707 | 95 (88–102) | 838 | 113 (105–120) |
| Asplenia/sickle cell disease | 2,545 | 141 (136–147) | 2,878 | 160 (154–166) | 1,089 | 146 (138–155) | 1,346 | 181 (171–191) |
| Other immunosuppression | 14,134 | 784 (771–797) | 32,537 | 1,805 (1,786–1,825) | 5,035 | 676 (658–695) | 7,663 | 1,029 (1,006–1,052) |
| Chronic kidney disease | 159,241 | 8,835 (8,793–8,876) | 159,379 | 8,842 (8,801–8,884) | 54,661 | 7,342 (7,283–7,401) | 54,774 | 7,357 (7,298–7,417) |
| Severe obesity (BMI ≥40 kg/m<sup>2</sup>) | 36,668 | 2,581 (2,555–2,607) | 36,668 | 2,581 (2,555–2,607) | 16,706 | 2,898 (2,855–2,942) | 16,706 | 2,898 (2,855–2,942) |
| **At-risk population** |  |  |  |  |  |  |  |  |
| At-risk population<sup>1</sup> | 450,601 | 24,999 (24,936–25,062) | 483,071 | 26,801 (26,736–26,865) | 175,420 | 23,562 (23,466–23,659) | 188,716 | 25,348 (25,249–25,447) |
| among those aged <70 years | 276,158 | 17,756 (17,696–17,816) | 303,528 | 19,516 (19,453–19,578) | 112,168 | 17,286 (17,194–17,378) | 123,038 | 18,961 (18,866–19,057) |
| among those aged ≥70 years | 174,443 | 70,579 (70,399–70,759) | 179,543 | 72,643 (72,466–72,818) | 63,252 | 66,157 (65,856–66,457) | 65,678 | 68,694 (68,399–68,988) |
| Multimorbidity<sup>2</sup> | 133,803 | 7,423 (7,385–7,462) | 155,802 | 8,644 (8,603–8,685) | 48,009 | 6,449 (6,393–6,505) | 56,827 | 7,633 (7,573–7,693) |
| **Cancer survivors** |  |  |  |  |  |  |  |  |
| Incident cancer in previous year | 8,343 | 463 (453–473) | 10,680 | 593 (581–604) | 2,936 | 394 (380–409) | 3,806 | 511 (495–528) |
| Incident cancer in previous 5 years | 31,195 | 1,731 (1,712–1,750) | 36,815 | 2,042 (2,022–2,063) | 11,708 | 1,573 (1,544–1,601) | 13,854 | 1,861 (1,830–1,892) |

95% CI, 95% confidence interval
1. The clinically vulnerable population comprised individuals with: any history of chronic respiratory disease other than asthma, heart disease, kidney disease, neurological conditions such as multiple sclerosis, diabetes mellitus; or with current asthma, severe obesity, or immunosuppression; assessed at each index date.
2. Multimorbidity was defined as more than one condition among the following domains: asthma or other chronic respiratory disease; chronic heart disease; chronic kidney disease; chronic liver disease; chronic neurological disease; diabetes; or immunosuppression (including individuals with dysplasia and organ transplant recipients).

### Sensitivity analyses

Age-standardisation did not alter 2019 at-risk prevalence estimates (not presented). When the study population was restricted to individuals active on 5 March 2019, at-risk prevalence estimates fell by less than 1% (**Supplementary Table 4**).

## Discussion

This paper describes the size and distribution of the population at risk of severe COVID-19 based on clinical records from a large, nationally representative cohort across the UK.

On 5 March 2019, 24·4% of the UK population were at higher risk than others of the same age due to underlying health conditions, including 8·3% of school-aged children, 19·6% of working-aged adults, and 66·2% of individuals aged ≥70 years. The commonest conditions were chronic kidney disease, diabetes and asthma. Multimorbidity was common at 7·1%. The size and regional distribution of the at-risk population was similar in 2014 and 2019, with lower prevalence in London and the South of England than Midlands or Northern regions. Separately, the 1·6% of the study population with a new diagnosis of cancer within the previous five years may also be at increased risk of severe COVID-19.^3^

Including all individuals aged ≥70 years, 18·5 million individuals in the UK would be considered moderate or high risk under current national guidance.^1^ This is higher than a previous estimate of 8.4 million comprising 7·2% of men and 7·5% of women aged 30–69 years, and 33% of men and 29% of women aged ≥70 years.^10^ Our estimates include additional conditions to cover the full national guidance.^1^ There were also differences in ascertainment: for example, our diabetes prevalence estimate for ages 30–69 in England was 7·0%, compared to 2·2% in the previous study^10^ and 6·9% in the national Quality Outcomes Framework (QOF).^18^ This may be due to increases in diagnoses and recording of diabetes over time in our more recent study period of March 2019 (rather than 1997–2017).^10^

Our at-risk prevalence estimates were slightly lower than GBD-based estimates that 29·1% of the UK population had at least one underlying health condition increasing COVID-19 risk, or 28·1% when restricted to the same set of conditions (by excluding cancers causing indirect immunosuppression and tuberculosis from GBD-based estimates).^7^ This did not appear to be due to under-estimation of clustering due to multimorbidity in the GBD-based study, as the 9·2% multimorbidity prevalence modelled was higher than we observed even when using linked secondary care records. The difference was greatest among older age groups, and our finding that 19·6% of working-aged adults (19–65 years) were at risk is broadly comparable to the GBD-based estimate (for the same conditions) of 22·8% among those aged 15–64 years.^7^

Our prevalence estimates are in line with national QOF diabetes and cancer monitoring, slightly higher than the more narrowly defined QOF chronic heart disease estimates,^18^ and consistent with previous UK studies of chronic kidney disease and asthma.^19, 20^ The five-year trends of increasing diabetes and decreasing asthma prevalence are consistent with directions of change in a previous study of asthma,^19^ and QOF, although 2014 QOF diabetes prevalence was slightly higher at 6·2% in 2013/14.^18^

Linked secondary care records in England increased the estimated size of the at-risk population only modestly, but the estimated prevalence of chronic liver disease in 2019 nearly doubled from 0·27% to 0·53%, and multimorbidity and chronic heart disease prevalence also increased. Our chronic liver disease prevalence estimate in England of 529/100,000 when supplemented with secondary care data is more consistent with previous national estimates of approximately 600/100,000 for the UK than our lower estimate using primary care data alone.^21^ Several studies of the associations between underlying health conditions and COVID-19 outcomes in England have used standalone primary care records to characterise underlying health conditions.^4, 16^ Such studies may under-ascertain chronic liver disease, heart disease and multimorbidity, and thus underestimate associations of these conditions with COVID-19 outcomes. If the risk of severe COVID-19 differs between underlying health conditions, then their differential under-ascertainment in primary health records may bias estimates of associations of underlying health conditions with COVID-19 outcomes.

Among women who were pregnant on 5 March 2014, 12·9% were at risk due to an underlying health condition, compared to a third of the pregnant women admitted to hospital with COVID-19.^22^ While the pregnancy register has high sensitivity for livebirths, pregnancy losses may be under-recorded.^14^ The 2·1% point prevalence estimate of pregnancy is perhaps low compared to a survey in which 591/5686 (10%) of women aged 16-44 years in Britain reported a pregnancy ending in the previous year, although these are not easily comparable.^23^ Caution is required in applying historical pregnancy estimates, as COVID-19 may affect family planning.

### Strengths and limitations

To our knowledge, these are the first prevalence estimates of the full population at risk of severe COVID-19 across the UK according to national guidelines. Strengths include the large, nationally representative cohort, risk group definitions with detailed ascertainment tailored to risk of COVID-19, and quantification of the value of linked secondary care records. To support policy and planning flexibly as evidence of the associations of different underlying conditions with COVID-19 outcomes evolves, we provide age-and region-stratified prevalence for each underlying condition separately, including separating asthma from other respiratory conditions.^16, 24^

A key limitation is that UK-wide estimates rely on primary care records, which may miss undiagnosed conditions and under-ascertain conditions diagnosed in secondary care. Our analysis including linked secondary care records in England suggests that estimates of the overall size of the at-risk population are robust, but that the prevalence of multimorbidity, chronic heart disease and liver disease may be underestimated from primary care records. There is likely under-ascertainment of immunosuppressing cancer treatments even using secondary care records, which could be on a scale similar to the 1·6% of the population newly diagnosed with cancer within the previous year. Second, the 2019 estimates did not include all regions in England. Although the dataset remained nationally representative in terms of age and sex in 2019, and prevalence estimates of individual conditions were consistent with expectations, suggesting that national 2019 estimates are representative, regionally-stratified estimates in 2019 are incomplete. Prevalence estimates from 2014 include all regions but are less up-to-date, and differences from 2019 may reflect changes in prevalence and recording of conditions, and the CPRD GOLD population over time. Third, inclusion of individuals active in the dataset at any point between 1 January and 5 March could have resulted in some under-estimation of point prevalence on 5 March: sensitivity analysis suggested this was minimal. Finally, we were able to describe pregnancy in 2014 only, and pregnancy prevalence may be under-estimated.

### Policy relevance

We estimate that current national guidance on COVID-19 risk groups encompasses 18·5 million individuals across the UK, a larger population than previously estimated.

Other studies have found that the health conditions in national COVID-19 guidance are indeed associated with increased risk of severe COVID-19, but to varying extents.^3, 4^ Older individuals are at higher risk of severe COVID-19 than younger, independent of underlying health conditions.^3, 4^

Implementation of public health measures such as influenza vaccination generally achieve higher uptake when targeted on the basis of age rather than health conditions.^25^ We found that 66·2% of individuals aged ≥70 years had at least one recorded underlying condition, suggesting that an age-based approach to COVID-19 vaccination could efficiently target individuals at highest risk. Age-based vaccination strategies may also be more feasible to implement in low-resource settings.

Our finding that 8·3% of school-aged children and 19·6% of working aged adults are in the at-risk population (as currently defined) emphasises the need to consider younger at-risk individuals in shielding guidance and when reopening schools and workplaces. The large number of children and younger adults with underlying conditions, who may nevertheless be at low absolute risk of severe COVID-19, supports vaccine strategies based on age- and condition-specific estimates of risk of severe COVID-19, rather than including individuals of any age with underlying conditions. We provide age-stratified prevalence for each condition to support effective vaccine resource allocation based on age and health conditions.

### Data sharing

The data used for this study were obtained from the Clinical Practice Research Datalink (CPRD). All CPRD data are available via an application to the Independent Scientific Advisory Committee (see https://www.cprd.com/Data-access). Data acquisition is associated with a fee and data protection requirements. This manuscript is supported by shared codelists used to define each condition, and estimates of the prevalence of the at-risk population and each underlying health condition, stratified by age and region (separately and combined) are available at https://doi.org/10.17037/DATA.00001833.

### Contributors

MR, JLB, RME, JLW and HIM conceived the study. HIM, DJG, JLW, HS, RME, IJD, NA, JLB, JAS, and LS designed the study. HIM, JLW, DJG, JKQ, HS, MP, CM, KEM, CTR, IJD, RM, AYSW designed the study variable definitions. JLW and DJG led the data extraction and data analysis with data analysis by HS, MP, CM, KEM, CTR, IJD, WM, AYSW, and HIM. All authors contributed to interpretation of results. HIM, JLW and HS drafted the manuscript to which all authors contributed, revised critically and approved. HIM is the guarantor. The corresponding author (HIM) attests that all listed authors meet authorship criteria and that no others meeting the criteria have been omitted.

#### Box 1

##### The COVID-19 moderate- and high-risk population defined in national guidance^1^ compared to the at-risk study population

| COVID-19 moderate- and high-risk population definitions in national guidance for England<sup>1</sup> | Study definition of the at-risk population |
| --- | --- |
| Anyone aged 70 years or older (regardless of medical conditions) is defined as at moderate risk.<br>Among individuals aged less than 70 years, people at moderate risk from coronavirus infection include people who:<br>• have a lung condition that's not severe (such as asthma, COPD, emphysema or bronchitis)<br>○ *high risk* have been told by a doctor they have a severe lung condition (such as cystic fibrosis, severe asthma or severe COPD)<br>• have heart disease (such as heart failure)<br>• have diabetes<br>• have chronic kidney disease<br>• have liver disease (such as hepatitis)<br>• have a condition affecting the brain or nerves (such as Parkinson's disease, motor neurone disease, multiple sclerosis or cerebral palsy)<br>• have a condition that means they have a high risk of getting infections<br>○ *high risk* have a condition that means they have a very high risk of getting infections (such as SCID or sickle cell)<br>○ *high risk* have blood or bone marrow cancer (such as leukaemia, lymphoma or myeloma)<br>• are taking medicine that can affect the immune system (such as low doses of steroids)<br>○ *high risk* are taking medicine that makes them much more likely to get infections (such as high doses of steroids or immunosuppressant medicine)<br>○ *high risk* have had an organ transplant<br>○ *high risk* are having chemotherapy or antibody treatment for cancer, including immunotherapy<br>○ *high risk* are having an intense course of radiotherapy (radical radiotherapy) for lung cancer<br>○ *high risk* are having targeted cancer treatments that can affect the immune system (such as protein kinase inhibitors or PARP inhibitors)<br>○ *high risk* have had a bone marrow or stem cell transplant in the past 6 months, or are still taking immunosuppressant medicine<br>• are very obese (a BMI of 40 or above) | The study definition of the at-risk population for all ages comprised individuals with any of the underlying health conditions in national guidance, incorporating high risk groups as indicated.<br>Detailed inclusion criteria for each condition were based on national guidance defining influenza clinical risk groups, to reflect risk of respiratory infection.<sup>26</sup> |
| • are pregnant<br>○ *high risk* have a serious heart condition and are pregnant | The prevalence of pregnancy, with and without any underlying health condition, was estimated separately from the general at-risk population. |

## Data Availability

The data used for this study were obtained from the Clinical Practice Research Datalink (CPRD). All CPRD data are available via an application to the Independent Scientific Advisory Committee (see https://www.cprd.com/Data-access). Data acquisition is associated with a fee and data protection requirements.
This manuscript is supported by shared codelists used to define each condition, and full estimates of the prevalence of the at-risk population and each underlying health condition, stratified by age and region (separately and combined) are provided online.

https://doi.org/10.17037/DATA.00001833

## References

1. NHS. People at higher risk from coronavirus 2020 [26/06/2020]. Available from: https://web.archive.org/web/20200716054208/https://www.nhs.uk/conditions/coronavirus-covid-19/people-at-higher-risk/whos-at-higher-risk-from-coronavirus/.

2. Docherty AB, Harrison EM, Green CA, Hardwick HE, Pius R, Norman L, et al. Features of 20 133 UK patients in hospital with covid-19 using the ISARIC WHO Clinical Characterisation Protocol: prospective observational cohort study. BMJ. 2020 May 22;369:m1985

3. Williamson EJ, Walker AJ, Bhaskaran K, Bacon S, Bates C, Morton CE, et al. OpenSAFELY: factors associated with COVID-19 death in 17 million patients. Nature. 2020 Jul 8

4. de Lusignan S, Dorward J, Correa A, Jones N, Akinyemi O, Amirthalingam G, et al. Risk factors for SARS-CoV-2 among patients in the Oxford Royal College of General Practitioners Research and Surveillance Centre primary care network: a cross-sectional study. Lancet Infect Dis. 2020

5. Jordan RE, Adab P. Who is most likely to be infected with SARS-CoV-2? Lancet Infect Dis. 2020 2020/05/15/

6. Crooke SN, Ovsyannikova IG, Poland GA, Kennedy RB. Immunosenescence and human vaccine immune responses. Immunity & ageing: I & A. 2019;16:25

7. Clark A, Jit M, Warren-Gash C, Guthrie B, Wang HHX, Mercer SW, et al. Global, regional, and national estimates of the population at increased risk of severe COVID-19 due to underlying health conditions in 2020: a modelling study. The Lancet Global health. 2020 Jun 15

8. Rezende LFM, Thome B, Schveitzer MC, Souza-Júnior PRB, Szwarcwald CL. Adults at high-risk of severe coronavirus disease-2019 (Covid-19) in Brazil. Revista de saude publica. 2020;54:50

9. Koma W, Neuman T, Claxton G, Rae M, Kates J, Michaud J. How many adults are at risk of serious illness if infected with Coronavirus? updated data. 2020 Available from: https://www.kff.org/coronavirus-covid-19/issue-brief/how-many-adults-are-at-risk-of-serious-illness-if-infected-with-coronavirus/.

10. Banerjee A, Pasea L, Harris S, Gonzalez-Izquierdo A, Torralbo A, Shallcross L, et al. Estimating excess 1-year mortality associated with the COVID-19 pandemic according to underlying conditions and age: a population-based cohort study. Lancet (London, England). 2020 May 30;395(10238):1715–25

11. Herrett E, Gallagher AM, Bhaskaran K, Forbes H, Mathur R, van Staa T, et al. Data Resource Profile: Clinical Practice Research Datalink (CPRD). Int J Epidemiol. 2015 Jun;44(3):827–36

12. Herrett E, Thomas SL, Schoonen WM, Smeeth L, Hall AJ. Validation and validity of diagnoses in the General Practice Research Database: a systematic review. British journal of clinical pharmacology. 2010 Jan;69(1):4–14

13. Herbert A, Wijlaars L, Zylbersztejn A, Cromwell D, Hardelid P. Data Resource Profile: Hospital Episode Statistics Admitted Patient Care (HES APC). Int J Epidemiol. 2017 Aug 1;46(4):1093-i

14. Minassian C, Williams R, Meeraus WH, Smeeth L, Campbell OMR, Thomas SL. Methods to generate and validate a Pregnancy Register in the UK Clinical Practice Research Datalink primary care database. Pharmacoepidemiol Drug Saf. 2019 Jul;28(7):923–33

15. Lewis JD, Bilker WB, Weinstein RB, Strom BL. The relationship between time since registration and measured incidence rates in the General Practice Research Database. Pharmacoepidemiol Drug Saf. 2005 Jul;14(7):443–51

16. Williamson E, Walker AJ, Bhaskaran KJ, Bacon S, Bates C, Morton CE, et al. [Preprint] OpenSAFELY: factors associated with COVID-19-related hospital death in the linked electronic health records of 17 million adult NHS patients. *medRxiv*. 2020:2020.05.06.20092999

17. Office for National Statistics. Population estimates for the UK, England and Wales, Scotland and Northern Ireland: mid-20192020. Available from: https://www.ons.gov.uk/peoplepopulationandcommunity/populationandmigration/populationestimates/bulletins/annualmidyearpopulationestimates/mid2019estimates.

18. Public Health England. Cardiovascular Disease, Diabetes and Kidney Disease 2020 [26/06/2020]. Available from: https://fingertips.phe.org.uk/profile-group/cardiovascular-disease-diabetes-kidney-disease.

19. Bloom CI, Saglani S, Feary J, Jarvis D, Quint JK. Changing prevalence of current asthma and inhaled corticosteroid treatment in the UK: population-based cohort 2006-2016. Eur Respir J. 2019 Apr;53(4)

20. McDonald HI. The epidemiology of infections among older people with diabetes mellitus and chronic kidney disease. London: London School of Hygiene & Tropical Medicine; 2015.

21. Pimpin L, Cortez-Pinto H, Negro F, Corbould E, Lazarus JV, Webber L, et al. Burden of liver disease in Europe: Epidemiology and analysis of risk factors to identify prevention policies. Journal of Hepatology. 2018 2018/09/01/;69(3):718–35

22. Knight M, Bunch K, Vousden N, Morris E, Simpson N, Gale C, et al. Characteristics and outcomes of pregnant women admitted to hospital with confirmed SARS-CoV-2 infection in UK: national population based cohort study. BMJ. 2020 Jun 8;369:m2107

23. Wellings K, Jones KG, Mercer CH, Tanton C, Clifton S, Datta J, et al. The prevalence of unplanned pregnancy and associated factors in Britain: findings from the third National Survey of Sexual Attitudes and Lifestyles (Natsal-3). Lancet (London, England). 2013 Nov 30;382(9907):1807–16

24. Halpin DMG, Faner R, Sibila O, Badia JR, Agusti A. Do chronic respiratory diseases or their treatment affect the risk of SARS-CoV-2 infection? The Lancet Respiratory Medicine. 2020 2020/05/01/;8(5):436–8

25. Public Health England. Surveillance of influenza and other respiratory viruses in the UK: Winter 2019 to 20202020. Available from: https://assets.publishing.service.gov.uk/government/uploads/system/uploads/attachment_data/file/895233/Surveillance_Influenza_and_other_respiratory_viruses_in_the_UK_2019_to_2020_FINAL.pdfW.

26. Public Health England. Immunisation against Infectious Disease (the Green Book). Chapter 19: Influenza. Available from: https://www.gov.uk/government/publications/influenza-the-green-book-chapter-19.

27. Nissen F, Morales DR, Mullerova H, Smeeth L, Douglas IJ, Quint JK. Validation of asthma recording in the Clinical Practice Research Datalink (CPRD). BMJ Open. 2017 Aug 11;7(8):e017474

28. Strongman H, Gadd S, Matthews A, Mansfield KE, Stanway S, Lyon AR, et al. Medium and long-term risks of specific cardiovascular diseases in survivors of 20 adult cancers: a population-based cohort study using multiple linked UK electronic health records databases. Lancet (London, England). 2019 Sep 21;394(10203):1041–54

29. Grint DJ, McDonald HI, Walker JL, Amirthalingam G, Andrews N, Thomas S. Safety of inadvertent administration of live zoster vaccine to immunosuppressed individuals in a UK-based observational cohort analysis. BMJ Open. 2020 Jan 29;10(1):e034886

